# Diagnostic performance of standard and inverted grey-scale CXR in detection of lung lesions in COVID-19 patients. A single institution study in the region of Abu Dhabi

**DOI:** 10.1101/2021.02.01.21250914

**Authors:** Abeer Ahmed AlHelali, Mohamed Ashfaque Kukkady, Ghufran Aref Saeed, Tamer Ibrahim Elholiby, Rabab Abdulrahman Al Mansoori, Luai A Ahmed

## Abstract

**Purpose:** To evaluate diagnostic performance of greyscale and inverted greyscale Chest X-ray (CXR) using Computed Tomography (CT) scan as a gold standard.

**Methods:** In this retrospective study, electronic medical records of 120 patients who had valid CXR and High-resolution CT (HRCT) within less than 24 hours after having a positive COVID-19 RT-PCR test during the period from May 19 ^th^ to May 23 ^rd^ 2020 in a single tertiary care center were reviewed.

PA chest radiographs were presented on 2 occasions to 5 radiologists to evaluate the role and appropriateness of greyscale and inverted greyscale chest radiographs (CXR). The images were viewed on high-specification viewing systems using a primary display monitors and compared it to computed tomography (CT) findings for screening and management of suspected or confirmed COVID-19 patients.

**Results:** Ninety-six (80%) patients had positive CT findings, 81 (67.5%) had positive grey scale CXR lesions, and 25 (20.8%) had better detection in the inverted grey scale CXR. The CXR sensitivity for COVID-19 pneumonia was 93.8% (95% CI (86.2% - 98.0%) and the specificity was 48.7% (95% CI (32.4% - 65.2%). The CXR sensitivity of detection of lung lesions was slightly higher in male (95.1% (95% CI (86.3% - 99.0%)) than female (90.0% (95% CI (68.3% - 98.8%)), while the specificity was 48.0% (95% CI (27.8% - 68.7%) and 50.0% (95% CI (23.0% - 77.0%) in males and females, respectively. However, no significant difference was detected in ROC area between men and women.

**Conclusions:** The sensitivity of detection of lung lesions of CXR was relatively high, particularly in men. The results of the study support the idea of considering conventional radiographs as an important diagnostic tool in suspected COVID-19 patients especially in healthcare facilities where there is no access to HRCT scans.

**Highlights:**

- CXR shows high sensitivity for detecting lung lesions in HRCT confirmed COVID-19 patients.
- Better detection of lesions was noted in the inverted grey scale CXR in (20.8%) of cases with positive findings in standard greyscale CXR.
- Conventional radiographs can be used as diagnostic tools in suspected COVID-19 patients especially in healthcare facilities where there is no access to HRCT scans.

## 1. Introduction

The pandemic caused by the novel coronavirus named COVID-19 or SARS-CoV-2, was first reported in China in December 2019 followed by widespread into all other countries in the world. Millions of confirmed cases have been found in the world [1]. COVID-19 clinical spectrum varies from asymptomatic or mild symptoms in most of the cases to severe acute respiratory syndrome (SARS), which may lead to death.

Most of the countries over the world are adopting molecular assay to screen for COVID-19 infection based on WHO guidelines [2]. Suspected COVID-19 patients are often seen in emergency department, trial to develop new strategies to diagnosed COVID-19 patients rapidly is the current challenge as the gold standard RT-PCR results need several hours, and have more than 5% false negative rates [3,4]. There are multiple typical features on chest HRCT which help in the treatment plan of suspected COVID-19 patients till the RT-PCR results are available such as bilateral, peripheral ground-glass opacities with or without consolidations predominantly affecting posterior segments [5]; however, the challenge of continuous infection control between the cases and the huge burden for the radiology departments should be taken into consideration [6].

Chest X-ray is widely available in emergency departments in particular, to complete the assessment of patients with respiratory symptoms or sepsis with minimal risk of cross infection as the surface of portable unit can be cleaned easily. Moreover, COVID-19 patients manifest with characteristic chest x-ray imaging features, which are helpful for the early detection of COVID-19 pneumonic changes as well as reflecting the severity of disease and following it up by comparing pre and post treatment changes [7]. However, the diagnostic performance of CXR in COVID-19 patients is not yet thoroughly investigated. This study aimed to evaluate diagnostic performance of greyscale and inverted greyscale CXR with CT scan as a gold standard in COVID-19 patients.

## 2. Materials and method

### 2.1. Study population

All suspected and confirmed COVID-19 patients admitted to the emergency department in a single tertiary care center in Abu Dhabi, UAE, who had RT-PCR test during the period from May 19 ^th^ to May 23 ^rd^ 2020 were eligible for this study. A total of 250 patients were admitted during the study period. Among them, 127 patients with negative RT-PCR results, no x-ray and HRCT of chest within 24 hours or had duplicate medical record numbers were excluded. A total of 120 COVID-19 positive patients who had chest X-ray and confirmatory HRCT chest within less than 24 hours from RT-PCR test results were included in the analysis.

The electronic medical records of these 120 patients were retrospectively reviewed. Demographic data on patients’ age and sex were extracted, and chest radiographs were reanalyzed.

Ethics approval was obtained from the local Research Ethics Committee (Abu Dhabi COVID19 Research Ethics Committee) Ref: DOH/CVDC/2020/1126.

### 2.2. Imaging technique and analysis

Five radiologists interpreted the chest radiographs during two reading sessions in standard view and inverted grey-scale view on primary display system. Primary display systems are used for interpretation of medical images, as in radiology. They have to meet strict performance criteria. On the other hand, secondary display systems are used by staff other than radiologists, usually after an interpretative report has been rendered. Chest X-ray was considered positive if alveolar, reticular opacity or both were found on CXR. All findings were compared to HRCT chest done within less than 24 hours. A high-resolution CT scan was performed in all patients with 64-slice multi-detector row CT scanners (VCT GE −64). Patients were scanned during breath hold craniocaudally in the supine position, from the lung apices to the costo-phrenic angles. The acquisition parameters were as follows: tube voltage 120 kV, tube current 100-600 mA, dose modulation by smart MA, pitch 1. The slice collimation 64x 0.625, slice width 0.625 x 0.625. The 1.25 mm or 2.5 mm thick images were reconstructed using lung window and a high-frequency reconstruction algorithm, then stored in the PACS system. Air purifier is fixed inside the scanner room and COVIDCIDE wipes are used to disinfect the scanner after completion of each examination. Sensitivity and specificity were used to assess accuracy based on presence or absence of lung changes. Patients with positive lung lesions manifested with patchy ground-glass opacity, linear opacities, or consolidations.

### 2.3. Statistical analysis

Descriptive statistics of patients’ demographic (age, sex) and imaging (X-ray, CT) characteristics are reported as means (standard deviation (SD)) and numbers and relative frequencies. The diagnostic performance of standard grey scale CXR was estimated using CT results as gold standard. Specificity, Sensitivity, positive predictive value, negative predictive value, and total accuracy of chest X-ray were estimated. The receiver-operating characteristic (ROC) curve analysis was used to calculate the area under the curve (AUC). The analysis was performed using STATA version 16.1 (Stata Corp, College Station, TX, USA), and p-value less than 0.05 defined statistical significance.

## 3. Results

### 3.1. Demographic and imaging data

The analysis included 120 confirmed cases of COVID-19. The majority of patients were male (86; 71.7%) with a mean age of 46.9 years (standard deviation 13.7 years) (Table 1). Ninety-six (80%) patients had positive CT findings, 81 (67.5%) had positive grey scale CXR lesions, and 25 (20.8%) had better detection in the inverted grey scale CXR (Table 1).

**Table 1:** Demographic characteristics and imaging findings of the patients.

|  | **N (%)** |
| --- | --- |
| N | 120 |
| Gender |  |
| Men | 86 (71.7) |
| women | 34 (28.3) |
| Age, years (mean (SD)) | 46.9 (13.7) |
| Age groups |  |
| 18 – 39 years | 35 (29.2) |
| 40 – 49 years | 42 (35.0) |
| 50 – 59 years | 23 (19.2) |
| 60+ years | 20 (16.7) |
| Grey scale CXR lesions |  |
| Yes | 81 (67.5) |
| No | 39 (32.5) |
| Inverted Grey scale lesions |  |
| Same findings | 95 (79.2) |
| Better detection | 25 (20.8) |
| CT findings |  |
| Positive | 96 (80.0) |
| Negative | 24 (20.0) |

### 3.2. X-ray features

Representative chest radiograph of COVID-19 pneumonia in a male patient (Figure 1) is showing subtle ground-glass opacity in CXR at the left lower zone with adjacent atelectatic changes (A) seen as well in the inverted greyscale image (B). Axial chest CT (C) and coronal view (D) confirmed the findings and are showing bilateral peripheral and central patchy ground-glass opacities.

**Figure 1:**
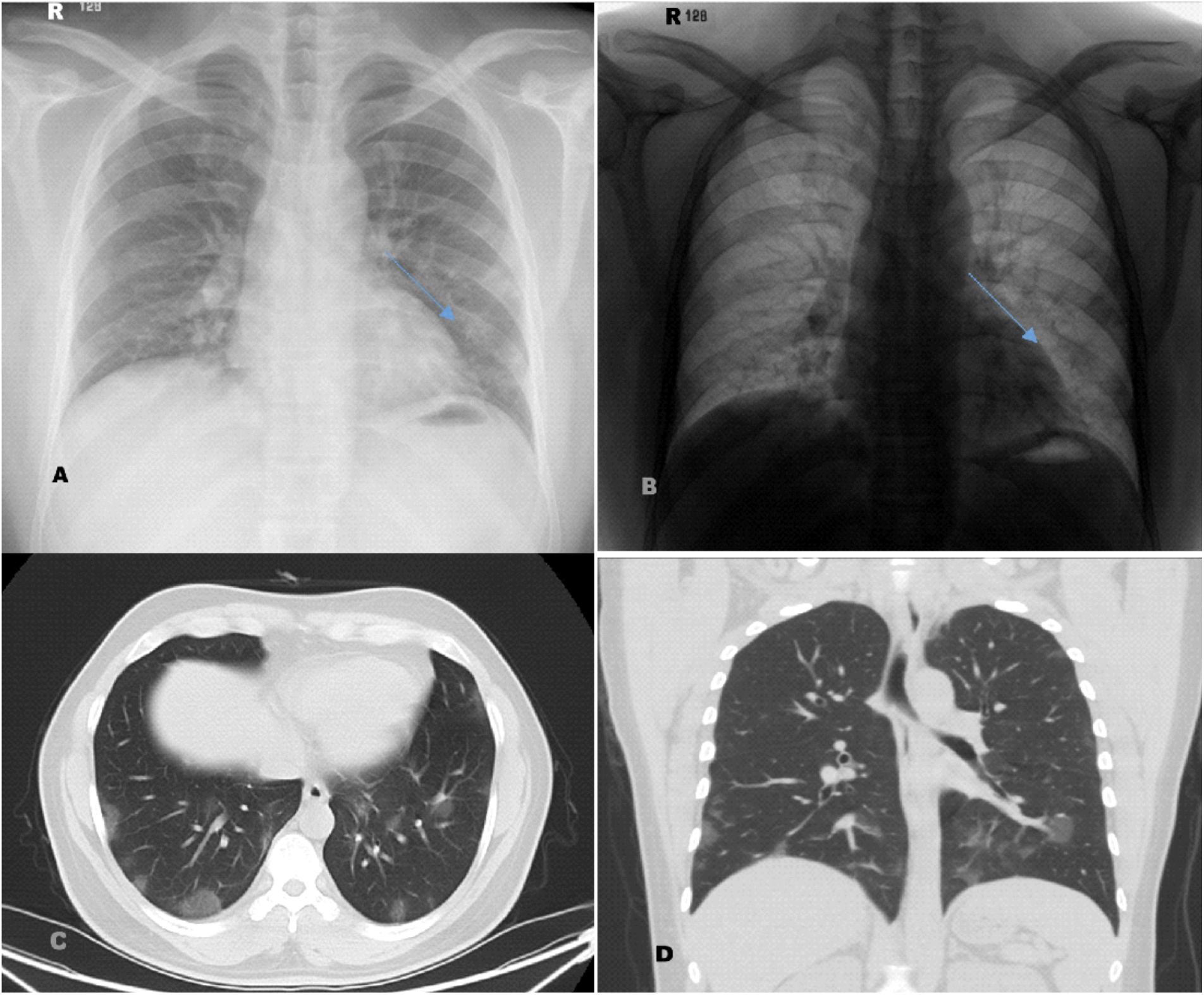
Representative chest radiograph (A) of COVID-19 pneumonia in a male patient, manifesting as subtle ground-glass opacity at the left lower zone with adjacent atelectatic changes, seen as well in inverted greyscale image (B). Axial chest CT (C) and coronal view (D) show bilateral peripheral and central patchy ground-glass opacities.

Another representative CXR of COVID-19 pneumonia (Figure 3) is showing a subtle ground-glass opacity at the base of both lungs with patchy opacity in the left middle zone (A), better seen in inverted greyscale image (B). Coronal chest CT images (C, D & E) confirmed the findings.

### 3.3. X-ray diagnostic performance

Using CT results as gold standard, the sensitivity and specificity of standard Greyscale CXR were 93.8% (95% CI (86.2% - 98.0%) and 48.7% (95% CI (32.4% - 65.2%), respectively (Table 2).Five cases out of 81 showed false positive results in CXR and 20 cases out of 39 showed false negative results. ROC area of standard grey scale CXR to CT was 0.713 (95% CI (0.629 - 0.796). Inverted grey scale showed similar findings compare to standard grey scale CXR in 95 patients (79.2%), however better detection of lung lesions was noted in 25 patients (20.8% of the cases). The CXR sensitivity of detection of lung lesions was better in male (95.1% (95% CI (86.3% - 99.0%)) than female (90.0% (95% CI (68.3% - 98.8%)), while the specificity was 48.0% (95% CI (27.8% −68.7%) and 50.0% (95% CI (23.0% - 77.0%) in males and females, respectively (Table 3). However, no significant difference was detected in ROC area between males and females (ROC area is 0. 715in male, 0.700 in female).

**Table 2:**
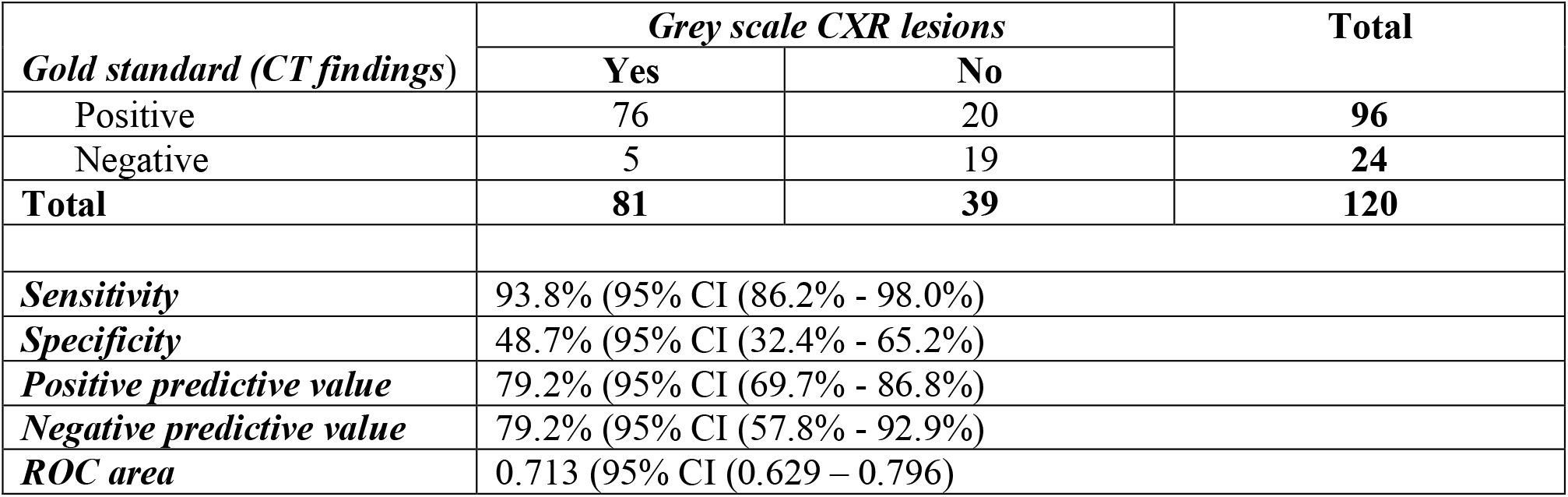
Diagnostic performance of Greyscale CXR lesions with CT as gold standard

**Table 3:** Diagnostic performance of Grey scale CXR lesions with CT as gold standard in men and women.

|  | **Men** |  |  | **Women** |  |  |
| --- | --- | --- | --- | --- | --- | --- |
| ***Gold standard (CT findings)*** | ***Grey scale CXR lesions*** |  | **Total** | ***Grey scale CXR lesions*** |  | **Total** |
|  | **Yes** | **No** |  | **Yes** | **No** |  |
| Positive | 58 | 13 | **71** | 18 | 7 | **25** |
| Negative | 3 | 12 | **15** | 2 | 7 | **9** |
| **Total** | **61** | **25** | **86** | **20** | **14** | **34** |
| ***Sensitivity*** | 95.1% (95% CI (86.3% - 99.0%)) |  |  | 90.0% (95% CI (68.3% - 98.8%)) |  |  |
| ***Specificity*** | 48.0% (95% CI (27.8% - 68.7%)) |  |  | 50.0% (95% CI (23.0% - 77.0%)) |  |  |
| ***Positive predictive value*** | 81.7% (95% CI (70.7% - 89.9%)) |  |  | 72.0% (95% CI (50.6% - 87.9%)) |  |  |
| ***Negative predictive value*** | 80.0% (95% CI (51.9% - 95.7%)) |  |  | 77.8% (95% CI (40.0% - 97.2%)) |  |  |
| ***ROC area*** * | 0.715 (95% CI (0.612 - 0.819)) |  |  | 0.700 (95% CI (0.548 - 0.852)) |  |  |

\* P-value for ROC differences between men and women was 0.869

## 4. Discussion

CXR showed high sensitivity for detecting lung lesions in HRCT confirmed COVID-19 patients. The performance was relatively higher in men than women. To the best of our knowledge, this is the first review and analysis of the diagnostic performance of standard and inverted grey-scale CXR in detection of lung lesions in COVID-19 patients with correlation to chest HRCT.

The results of the study support the idea of considering conventional radiographs as an important diagnostic test in suspected COVID-19 patients. The burden imposed by the COVID-19 pandemic on healthcare institutions highlighted the need to use a fast and available tool like chest x-ray to prioritize patients’ management and predict results. Therefore, CXR can play an important role in the detection of lung changes, while chest HRCT can have a complementary role in evaluating disease severity, progression and complications [9,10]. Prior studies on the pattern and frequency of CXR and CT opacities in COVID-19 positive patients have demonstrated that opacities are typically peripheral, basilar and bilateral in distribution predominantly affecting right lower lobe, especially on early stages of the disease [8,11,12,13]. Studies have reported that 33% to 86% of COVID-19 chest CTs showed peripheral lung distribution [6, 14]. On the other hand, chest x-ray may in many cases show unilateral air space opacities involving a single lobe, and this may still raise the possibility of COVID-19 pneumonia as it is challenging to identify all pneumonic changes on a limited CXR. In such cases, HRCT of the chest can help and may show the typical bilateral peripheral pneumonic changes as in (Fig. 1) The high sensitivity found in our study indicate that CXR is highly helpful in identifying such lesions in COVID-19 positive patients. The false positive CXR findings could be attributed to the overlapping of soft tissue which give ground glass opacity mimicking COVID-19 changes or old atelectasis.

However, CXR findings in COVID-19 patients might not be specific, and can be seen with other infections, including influenza, SARS, MERS and H1N1 [11,15].

Our findings showed a relatively lower specificity levels of CXR compared to HRCT as gold standard. Although, COVID-19 is considered a serious disease and high specificity would be preferred to avoid making false negative diagnosis, the low specificity found in this study could be attributed to mild cases not developing lung lesions that could be detected by CXR [16]. Moreover, the United Arab Emirates is undertaking an intensified testing campaign for COVID-19 accompanied by strong awareness program with early diagnosis and management of the cases. Therefore, many early detected positive COVID-19 cases might need time to develop lung manifestations which can be detected by CXR. A repeated CXR would be warranted in suspected patients.

The study had the advantage of having chest HRCT within less than 24 hours of performing portable chest x-ray for all COVID-19 positive patients and it also checked for the effect of using inverted greyscale images compared to standard grey scale CXR images. Interestingly, the inverted greyscale CXR showed better detection of lesions in 20.8% of the cases (fig. 2) than standard greyscale CXR.

**Figure 2:**
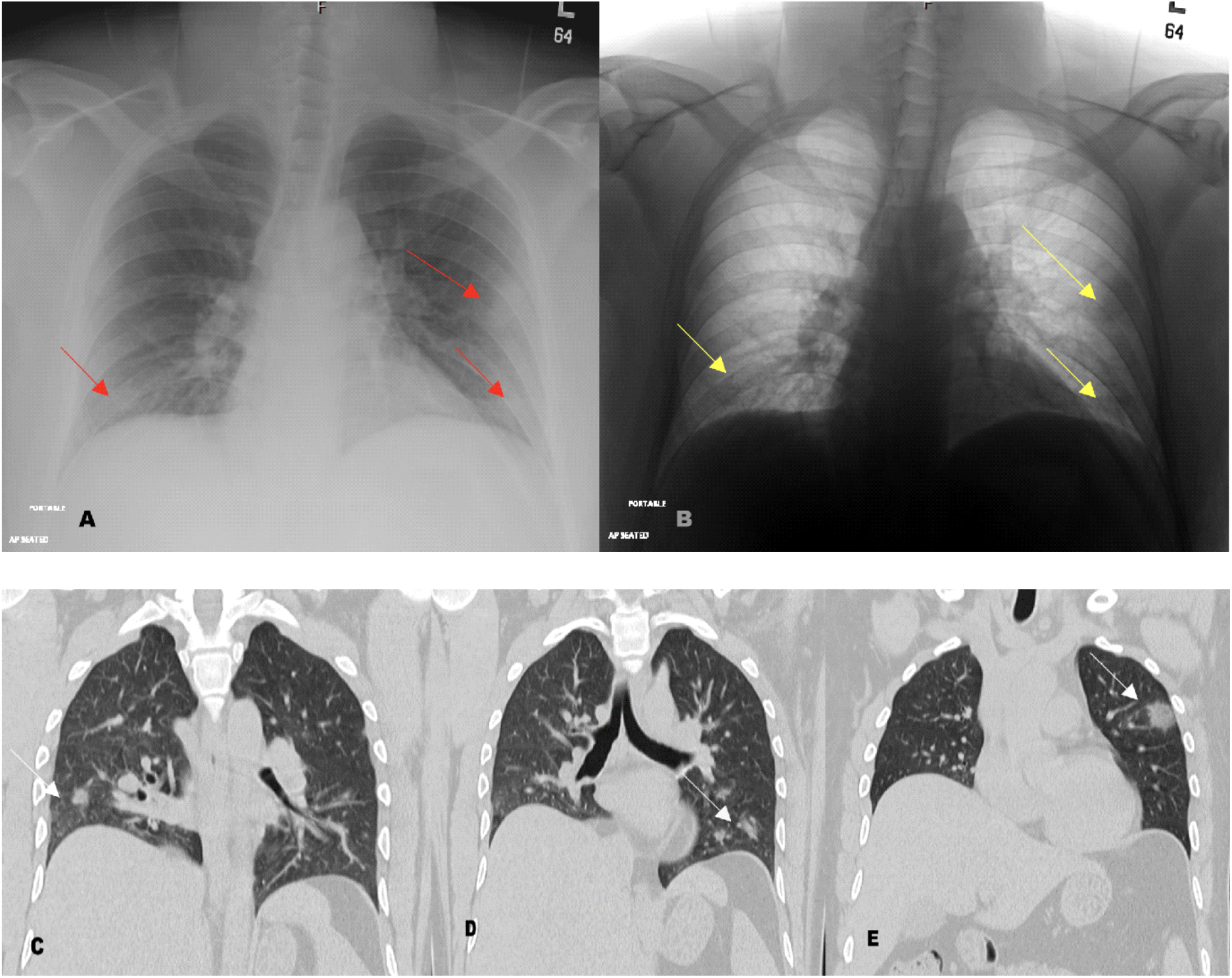
Representative chest radiograph (A) of COVID-19 pneumonia in a male patient, manifesting as subtle ground-glass opacity at the base of both lungs with patchy opacity in the left middle zone, better seen in inverted greyscale image (B). Coronal chest CT images (C, D&E) show patchy consolidative lesions (arrows) bilaterally in peripheral zones.

## Conclusions

The sensitivity of detection of lung lesions of CXR was relatively high, particularly in men. The results of the study support the idea of considering conventional radiographs with standard and inverted greyscale images as an important diagnostic tool in suspected COVID-19 patients at the emergency departments, especially in healthcare facilities where access to HRCT scan does not exist.

## Data Availability

Available data

## Abbreviations

CXR: Chest X-ray
CT: Computed Tomography
COVID-19: Corona virus disease 2019
HRCT: High-resolution CT
SARS-CoV-2: Severe Acute Respiratory Syndrome Corona-Virus 2
WHO: World Health Organization
RT-PCR: Reverse Transcriptase-Polymerase Chain Reaction
MERS: Middle East Respiratory Syndrome
H1N1: subtype of influenza A virus

## CRediT authorship contribution statement

Abeer Ahmed AlHelali: Conceptualization, Data collection, Formal Analysis, Writing - original draft, Writing - review & editing, Supervision, Project administration.

Mohamed Ashfaque Kukkady: Data collection, Formal analysis, Methodology, Writing - review & editing Supervision.

Ghufran Aref Saeed: Data collection, Formal analysis, Writing - review & editing.

Tamer Ibrahim Elholiby: Data collection, Methodology, Writing - review & editing.

Rabab Abdulrahman Al Mansoori: Data collection, Methodology, Writing - review & editing.

Luai Ahmed: Methodology, Formal analysis, Writing - review & editing.

## Acknowledgements

We would like to thank Mr.George Roy senior CT technician for providing technical information about chest HRCT, all the radiographers and CT technicians that have dedicated their effort and time to perform thousands of studies for COVID 19 patients in ED and in isolation wards and the radiologist staff at Sheikh Khalifa Medical City for their strong performance during this pandemic crisis to facilitate the speed of patient care.

